# Evaluation of Alinity m CMV assay performance for detecting CMV in plasma, cerebrospinal fluid, and bronchoalveolar lavage specimens

**DOI:** 10.1101/2023.11.29.23299213

**Authors:** Joshua Kostera, Mark Hubbard, Dillon Jackson, Rachael M. Liesman

**Author notes:** **Corresponding author:** Rachael Liesman, 8701 W Watertown Plank Road, Milwaukee WI 53226. **Funding** This study was funded by Molecular Diagnostics of Abbott.

## Abstract

**Introduction:** Rapid and accurate detection of cytomegalovirus (CMV) infection and reactivation is crucial to preventing adverse outcomes in immunocompromised individuals, especially in transplant recipients. Current PCR-based assays were developed for use with plasma specimens, but CMV is present in bronchoalveolar lavage fluid (BAL) in immunocompromised patients with CMV pneumonia and has also been detected in cerebrospinal fluid (CSF).

**Objectives:** We evaluated the performance of the Abbott Alinity m CMV assay compared to the Abbott RealTi*m*e CMV assay for quantification of CMV in plasma, BAL, and CSF specimens.

**Methods:** Analytical performance, including linearity, precision, and limit of detection, was established for plasma, BAL, and CSF specimen types using reference CMV control material (SeraCare). To evaluate clinical performance, 190 plasma specimens, 78 BAL specimens, and 26 CSF specimens were tested with the Alinity m assay and compared to the RealTi*m*e CMV assay.

**Results:** The Alinity m CMV assay showed high precision (SD <0.01 to 0.13) within the quantifiable range (1.49-8.00 log10 IU/mL) for all 3 specimen types, with strong clinical correlation to the RealTi*m*e CMV assay results (r^2^=0.9779 for plasma, r^2^=0.9373 for BAL, r^2^=0.9889 for CSF).

**Conclusion:** The Alinity m CMV assay may be useful for rapid detection of CMV in plasma, BAL, and CSF specimens.

## Introduction

Cytomegalovirus is highly prevalent worldwide(1). After initial infection, CMV becomes latent and can be reactivated in response to stress or suppression of the immune response (2). In immunocompetent individuals, primary CMV infection may be asymptomatic or mildly symptomatic; however, infection or reactivation of latent CMV in immunocompromised individuals is associated with severe to fatal outcomes(2-4). Individuals with hematopoietic stem cell or solid organ transplants are at high risk of graft rejection or loss, multi-organ failure, and death as a consequence of CMV reactivation(5). Ongoing monitoring of CMV DNA load in transplant recipients is therefore essential to prevent CMV-related adverse outcomes(6).

Highly sensitive and specific nucleic acid amplification tests (NAATs) for quantifying CMV DNA in plasma have been developed for the detection of CMV reactivation and response to anti-CMV treatment(7).CMV can also be detected in bronchoalveolar lavage (BAL) specimens from transplant recipients and immunocompromised patients with suspected CMV pneumonia(8-11). CMV infection of the central nervous system may occur in severely immunosuppressed patients, such as those with advanced HIV infection, and CMV may be detectable in the cerebrospinal fluid (CSF)(6, 12). Here, we evaluated the performance of the Alinity m CMV assay compared to the Abbott RealTi*m*e CMV assay for quantification of CMV viral load in plasma specimens and detection of CMV in CSF and BAL specimens.

## Materials and Methods

### Specimens

Deidentified remnant specimens (plasma, CSF, and BAL) from patients were utilized to assess the performance of the Alinity m CMV assay. All assays were run in the central laboratory of a large urban tertiary care hospital. The study was determined to be exempt by The University of Kansas Health System Institutional Review Board.

### Assay platforms

The Alinity m CMV assay is a real-time PCR-based test that targets regions within the UL34 and UL80.5 genes to detect and quantify CMV DNA in human plasma, with a reported quantitative range of 1.48 to 8.00 log10 IU/mL. The RealTi*m*e CMV assay is performed on the *m*2000 platform, consisting of the *m*2000*sp* for sample preparation and *m*2000*rt* for amplification and detection. The RealTi*m*e CMV assay assay has the same target region as the Alinity m CMV assay with a quantitative range of 1.70 to 8.19 log10 IU/mL.

### Analytical evaluation

To assess limit of detection (LoD) in plasma specimens, the ExactDX (EDX) CMV LoD panel (Exact Diagnostics, Fort Worth, TX) was tested at 1.65 log10 IU/mL and diluted in negative plasma to 1.48 log10 IU/mL; each level was tested in replicates of 20 across 5 days. The LoD was assessed in BAL and CSF by diluting the EDX panel to 2.00, 1.70, and 1.48 log10 IU/mL in negative matrix and each level was tested in replicates of 15. The lowest concentration with 100% qualitative detection was determined as the LoD. Precision for plasma specimens was assessed with EDX material at high (5.70 log10 IU/mL) and low (3.0 log10 IU/mL) concentrations tested in triplicate daily for 6 days. Linearity for plasma specimens was assessed with the EDX CMV panel, consisting of 6 levels (2.30, 2.60, 3.60, 4.60, 5.60, and 6.60 log10 IU/mL) tested in duplicate daily over 4 days. To assess linearity and precision of the Alinity m CMV assay in BAL and CSF specimens, CMV-negative pooled BAL or CSF specimens were spiked with a CMV-positive plasma sample at final concentrations of 2.60, 4.60, and 6.60 log10 IU/mL and tested in triplicate daily for 5 days. External Alinity CMV quality control material at high and low concentrations were also tested daily.

### Clinical specimen evaluation

To assess accuracy, a total of 190 deidentified remnant plasma specimens initially tested on the RealTi*m*e CMV assay as part of routine clinical care were tested on the Alinity m CMV assay. Specimens were stored refrigerated (2-8°C) for ≤48 hours (n=81) or frozen (-20°C; n=109) prior to testing. Additionally, 78 BAL specimens and 26 CSF specimens initially tested using the RealTi*m*e CMV assay were tested on the Alinity m CMV assay. Specimens were stored refrigerated (2-8°C) for ≤48 hours (BAL n=14; CSF n=7) or frozen (-20°C; BAL n=64; CSF n=19) prior to testing. Because all clinical CSF specimens were negative for CMV, positive CSF specimens were generated by spiking negative pooled CSF with a known CMV-positive plasma specimen (7.1 log10 IU/mL), which was then serially diluted to the analytical measurement range of 1.6 to 6.6 log 10 IU/mL. The dilutions were tested on both the RealTi*m*e CMV assay and Alinity m CMV assay.

### Statistical analysis

All analyses were performed using PC SAS (version 9.4) software (SAS, Cary, NC, USA). The percent positive agreement (PPA), negative percent agreement (NPA), and overall percent agreement (OPA) were calculated to determine qualitative method agreement. Variability is expressed as standard deviation (SD) and percent coefficient of variation (%CV). Correlation analysis was performed only on specimens with quantifiable results and was evaluated using by Deming regression analysis and Bland-Altman plots.

## Results

### Analytical performance

The Alinity m CMV assay was determined to be highly reproducible across all 3 specimen types (Table 1). In plasma, the standard deviation (SD) range was 0.07-0.11, inter-assay coefficients of variation (%CV) ranged from 1.27% to 3.45%, and intra-assay %CV from 0.84% to 2.32%. In BAL, SD ranged from 0.06-0.13 and inter- and intra-assay %CV ranged from 1.38% to 4.74% and 1.17% to 3.76%, respectively. In CSF, SD was below 0.1 and both inter- and intra-assay %CVs ranged from 0.71% to 2.12%. The Alinity m CMV assay demonstrated excellent linearity in plasma across the quantifiable range (1.49 and 8.00 log10 IU/mL), consistent with the assay package insert (Figure 1A). Linearity was also demonstrated in BAL and CSF specimens across the analytical measurement range (Figure 1B and 1C, respectively). The LoD for plasma, BAL, and CSF specimen types was determined to be 1.48 log10 IU/mL (30 IU/mL) for all matrices. Trend analysis of the high EQC material from 2 reagent lots by Levey-Jennings plot demonstrated 2 points outside of ±2SD acceptable range and no points oustide of ±3SD acceptable range across 29 non-consecutive days of testing (Figure 2A). A similar analysis of the low EQC material showed 1 point outside of ±2SD and no points outside of ±3SD acceptable ranges (Figure 2B).

**Table 1.** Precision of the Alinity m CMV Assay.

|  | Level<br>(log10<br>IU/mL) | Mean<br>(log10<br>IU/mL) | SD<br>(log10<br>IU/mL) | %<br><2SD | Inter-assay<br>%CV | Intra-assay<br>%CV |
| --- | --- | --- | --- | --- | --- | --- |
| EQC | N/A | 5.95 | 0.08 | 100 | 1.31 | 1.09 |
|  | N/A | 3.18 | 0.09 | 100 | 2.91 | 2.32 |
| Plasma | 5.70 | 5.68 | 0.07 | 100 | 1.27 | 0.84 |
|  | 3.00 | 3.05 | 0.11 | 100 | 3.45 | 2.24 |
| BAL | 6.60 | 6.33 | 0.13 | 100 | 2.04 | 1.32 |
|  | 4.60 | 4.59 | 0.06 | 100 | 1.38 | 1.17 |
|  | 2.60 | 2.58 | 0.12 | 93 | 4.74 | 3.76 |
| CSF | 6.60 | 6.59 | 0.09 | 100 | 1.32 | 1.02 |
|  | 4.60 | 4.81 | 0.07 | 93 | 1.44 | 0.71 |
|  | 2.60 | 2.75 | 0.06 | 100 | 2.12 | 1.87 |

BAL, bronchoalveolar lavage; CSF, cerebrospinal fluid; CV coefficient of variation; N/A, not available; SD, standard deviation; EQC, external kit quality control material.

**Figure 1.**
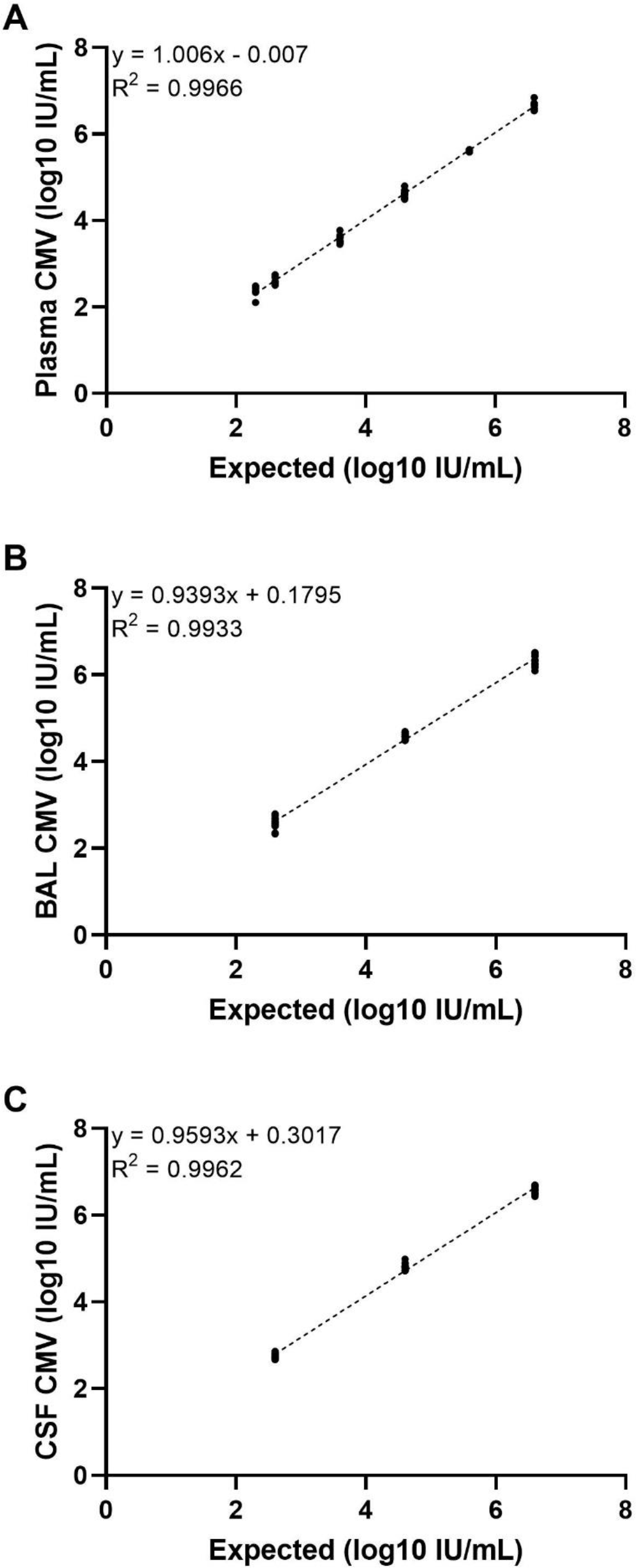
Alinity m CMV assay linearity for dilution series in (A) plasma, (B) BAL, and (C) CSF.

**Figure 2.**
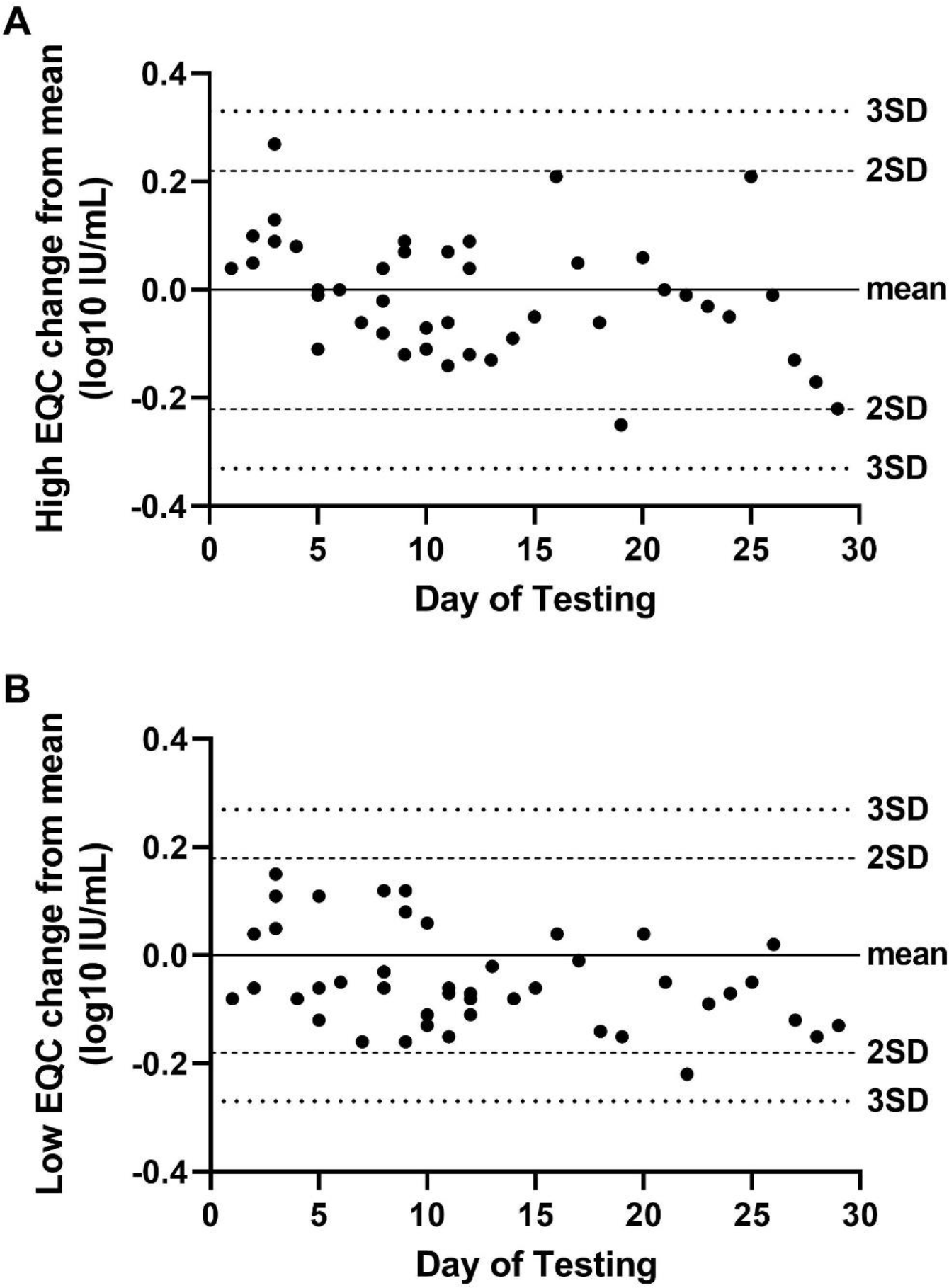
Levey-Jennings plots of (A) high and (B) low external QC. The dashed line represents the 2SD acceptable range and the dotted line represents the 3SD acceptable range.

### Clinical performance

One hundred and ninety plasma specimens were included in this study and qualitative positive, negative, and overall agreement rates were 95%, 89%, and 93%, respectively (Table 2). Fourteen specimens were discordant, with 8 specimens detected only by the Alinity m assay and 6 specimens detected only by the RealTi*m*e assay. All 14 discordant plasma specimens were detected below the limit of quantification of the respective assay (<50 IU/mL for the RealTi*m*e assay; <30 IU/mL for the Alinity m assay), consistent with detection variability at the LoD. A total of 97 specimens were within the quantifiable range of both assays and were used to assess clinical correlation between the assays. Deming regression analysis demonstrated high correlation between the 2 assays (R^2^ = 0.9779; slope = 1.0032) (Figure 3A). A Bland-Atlman plot was used to further analyze correlation between the methods (Figure 3B). Overall bias was -0.06 log10 IU/mL, with only 1 specimen demonstrating a bias >0.5 log10 IU/mL.

**Table 2.** Agreement Between RealTi*m*e CMV Assay and Alinity m CMV Assay for Plasma, BAL, and CSF.

| Plasma Specimens<br>(N=190) |  | RealTime |  | Positive Agreement | Negative Agreement | Overall Agreement |
| --- | --- | --- | --- | --- | --- | --- |
|  |  | Detected | Not Detected |  |  |  |
| Alinity m | Detected | 108 | 8 | 95% | 89% | 93% |
|  | Not Detected | 6 | 68 |  |  |  |
| BAL Specimens<br>(N=78) |  | RealTime |  | Positive Agreement | Negative Agreement | Overall Agreement |
|  |  | Detected | Not Detected |  |  |  |
| Alinity m | Detected | 48 | 4 | 100% | 87% | 95% |
|  | Not Detected | 0 | 26 |  |  |  |
| CSF Specimens<br>(N=26) |  | RealTime |  | Positive Agreement | Negative Agreement | Overall Agreement |
|  |  | Detected | Not Detected |  |  |  |
| Alinity m | Detected | 6 | 0 | 100% | 100% | 100% |
|  | Not Detected | 0 | 20 |  |  |  |

**Figure 3.**
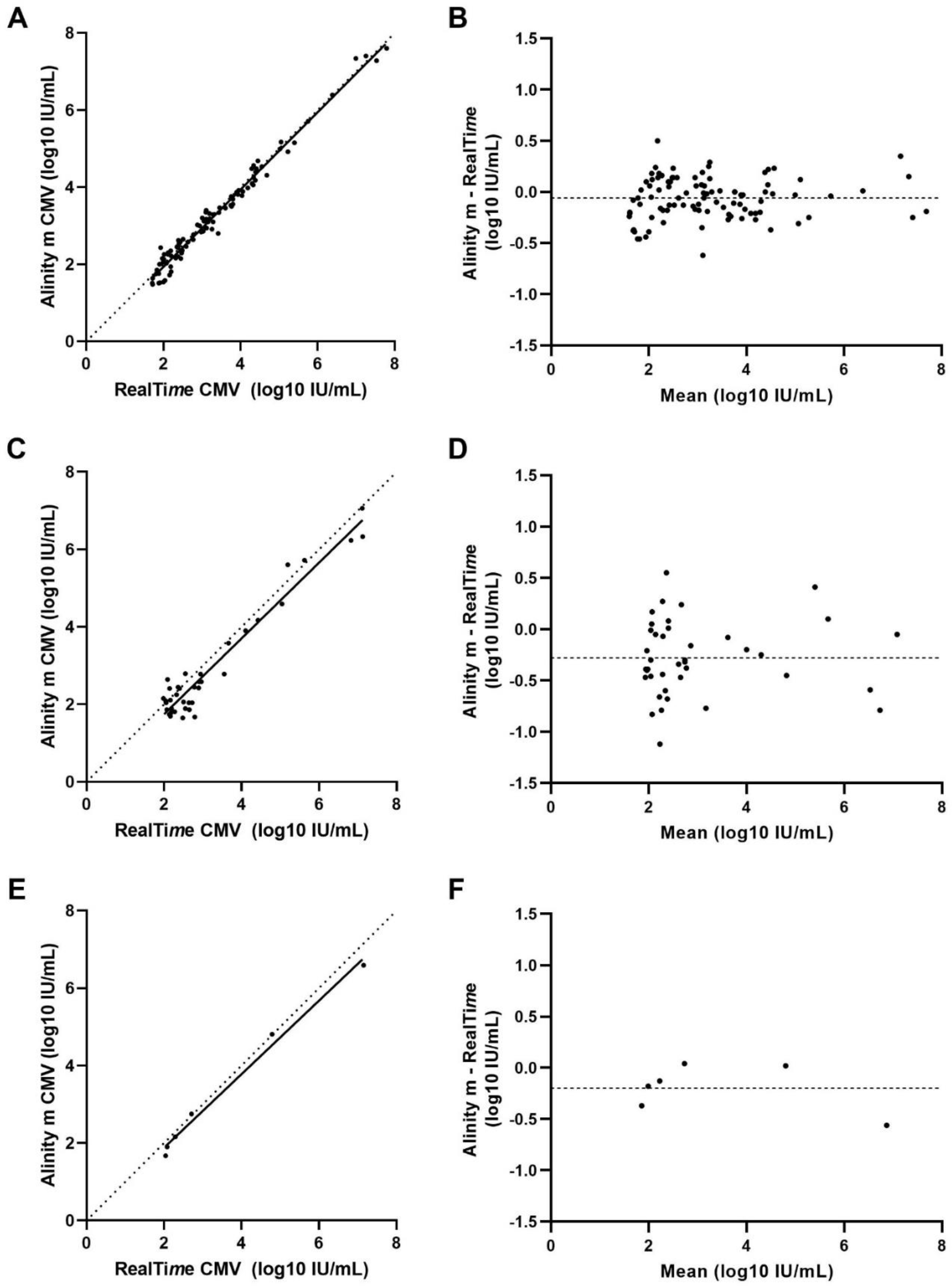
Comparison of Alinity m CMV and RealTi*m*e CMV assay performance in plasma, BAL, and CSF. (A) Deming regression of 97 plasma specimens within the quantifiable range of both assays and (B) Bland-Altman plot of bias. (C) Deming regression of 40 bronchoalveolar lavage (BAL) specimens within the quantifiable range of both assays and (D) Bland-Altman plot of bias. (E) Deming regression of 6 contrived CSF specimens within the quantifiable range of both assays and (F) Bland-Altman plot of bias. The dashed line in all Blant-Altman plots represents the average bias.

Seventy-eight BAL specimens were assessed for qualitative accuracy, with positive, negative, and overall agreement rates of 100%, 87%, and 95%, respectively (Table 2). Four total specimens were discordant, which were detected by the Alinity m assay below the quantifiable range (<30 IU/mL). A total of 40 specimens were within the quantifiable range of both assays. Deming regression analysis showed high correlation between the assays (R^2^=0.9373; slope 0.9781) (Figure 3C). Bland-Altman analysis demonstrated an overall bias of -0.28 log10 IU/mL (Figure 3D). Ten specimens (25%) had quantitative values with a bias >0.5 log10 IU/mL, with 9 specimens resulting in a >0.5 log10 IU/mL higher CMV DNA load from the RealTi*m*e assay and 1 specimen resulting in a >0.5 log10 IU/mL higher CMV DNA load when tested with the Alinity m assay.

All negative and contrived positive CSF specimens were concordant between the RealTi*m*e and Alinity m CMV assays (Table 2). For the 6 contrived positive specimens, Deming regression analysis showed high correlation (R^2^=0.9889; slope 0.9489) between the 2 assays (Figure 3E). Overall bias was -0.20 log10 IU/mL and 1 specimen demonstrated bias >0.5 log10 IU/mL (Figure 3F).

## Discussion

In this study, we assessed the analytical performance of the Alinity m CMV assay for quantitation of CMV viral load in plasma, CSF, and BAL specimens as compared to the RealTi*m*e CMV assay. Overall, the Alinity m assay demonstrated excellent linearity, precision, and clinical correlation across all 3 specimen types. Additionally, the LoD determined in this study for plasma samples (30 IU/mL) was consistent with the manufacturer’s package insert, and an equivalent LoD was also established for BAL and CSF specimens, which were not assessed by the manufacturer.

Our study findings are consistent with previous studies that have demonstrated the detection of CMV in BAL or bronchial brush specimens with PCR-based tests to diagnose CMV pneumonia in immunocompromised patients, patients with hematologic malignancies, and after transplantation(8, 13, 14). Previous studies have reported difficulty in defining quantitative cut-offs for CMV viral load in BAL specimens associated with CMV pneumonia(10). A recent prospective study identified a CMV load of 831 IU/mL as the diagnostic cut-off for CMV pneumonia; however, the authors recommended NAAT as a complementary test to bronchoscopy for definitive diagnosis(11). Further studies are needed to establish clinical decision points for CMV viral load values. Although we assessed the quantitative performance of this assay in CSF and BAL specimens, we report CMV testing from these specimens qualitatively.

In our comparative analysis, we found high percent agreement and correlation of results between Alinity m CMV and the RealTi*m*e CMV assay for all 3 specimen types. Of 294 total specimens assessed in this study, 18 (6%) were considered discordant. Most discordant samples were detected only by the Alinity m assay (n=12), which may be due to the lower LoD of the Alinity m assay as compared to the RealTi*m*e assay. Further, all discordant samples were detected below the limit of quantification, consistent with variable detection of viral DNA at the LoD. Further, no significant bias was demonstrated when comparing plasma specimens tested on both assays, which allows for a change of method without the need for re-baselining of patients. Greater bias was demonstrated when assessing clinical correlation of CMV DNA load in BAL specimens suggesting a potential need for re-baselining patients or reestablishing cut-off values for laboratories reporting quantitative results in BAL specimens.

In our single-center study, only a limited number of clinical CSF specimens were available for testing, and all were found to be negative for CMV. Larger studies are needed to establish the clinical performance of the Alinity m CMV assay with CSF specimens.

In conclusion, we found that the Alinity m CMV assay demonstrated high precision and accuracy for the quantitation of CMV DNA in both contrived panels and clinical specimens. The Alinity m platform allows rapid reporting of test results, which may improve turnaround time allowing for optimized detection and monitoring of CMV in at-risk patients.

## Data Availability

All data produced in the present study are available upon reasonable request to the authors

## Acknowledgements

This study was funded by Molecular Diagnostics of Abbott. Stacey Tobin, PhD, provided editorial support for manuscript preparation, with compensation from Abbott Laboratories.

